# Symptoms and severity in vaccinated and unvaccinated patients hospitalised with SARS-CoV-2 delta (B.1.617.2) variant infection

**DOI:** 10.1101/2022.02.08.22270506

**Authors:** Olivier Epaulard, Sophie Abgrall, Maeva Lefebvre, Jean-François Faucher, Jocelyn Michon, Emilia Frentiu, Cécile Janssen, Gabrielle Charbonnier, Audrey Fresse, Simon Laurent, Lena Sandjakian, Pierre Casez, Aba Mahamat, Guillaume Béraud, the Groupe Vaccination-Prévention, Société de Pathologie Infectieuse de Langue Française

**Affiliations:** Infectious diseases department, Centre Hospitalier Universitaire Grenoble-Alpes, Grenoble, France; Groupe de Recherche en Infectiologie Clinique, CIC-1406, Inserm-CHUGA-UGA, Grenoble, France; Service de Médecine Interne, Hôpital Antoine Béclère APHP,, Clamart; Université Paris-Saclay, UVSQ, INSERM U1018, CESP, Le Kremlin-Bicêtre, France; Infectious Diseases Department, Centre for Prevention of Infectious and Transmissible Diseases, CHU Nantes and INSERM UIC 1413 Nantes University, Nantes, France; CHU Limoges, Department of Infectious Diseases and Tropical Medicine, Limoges, France; Inserm U1094, IRD U270, Univ. Limoges, CHU Limoges, EpiMaCT; Department of Infectious diseases, University Hospital of Caen, Caen, France; Infectious diseases department, Centre Hospitalier Universitaire de Nancy, Nancy, France; Infectious Disease Unit, Centre Hospitalier Annecy Genevois, Annecy, France; Pharmacovigilance, Centre Hospitalier Universitaire de Nancy, Nancy, France; Regional Centre for Prevention of Infectious Diseases and Healthcare-Associated Infections, General Hospital of Ajaccio; Ajaccio, France; Department of Internal Medicine and Infectious Diseases, University Hospital of Poitiers; Poitiers, France

## Abstract

**Background:** The diffusion of the SARS-CoV-2 delta (B.1.617.2) variant and the waning of immune response after primary Covid-19 vaccination favoured the breakthrough SARS-CoV-2 infections in vaccinated subjects. To assess the impact of vaccination, we determined the severity of infection in hospitalised patients according to vaccine status.

**Methods:** We retrospectively analysed data from patients hospitalised in 10 centres with a SARS-CoV-2 infection (delta variant) from July to November 2021: i) all patients who had completed their primary vaccination at least 14 days before hospital admission; and ii) the same number of completely unvaccinated patients. We assessed the impact of vaccination and other risk factors through logistic regression.

**Findings:** We included 955 patients (474 vaccinated and 481 unvaccinated). Vaccinated patients were significantly older, more frequently males, and with more comorbidities. They were less often admitted for Covid-19 (59·3% vs. 75·1%, p<0·001), showed fewer lung lesions, and required oxygen less frequently (57·5% vs. 73·0%, p<0·001), at a lower flow (3·0 vs. 6·0 L/min, p<0·001), and for a shorter duration (3 vs. 6 days, p<0·001). They less frequently required intensive care unit admission (16·2 % vs. 36·0 %, p<0·001). Mortality at day 28 was not different between the two groups (16·7% vs. 12.2%, p=0·075), but multivariate logistic regression showed that vaccination significantly decreased the risk of negative outcomes, including mortality, even when considering older patients, and those with comorbidities.

**Conclusions:** Among patients hospitalised with a delta variant SARS-CoV-2 infection, vaccination was associated with less severe forms, even in the presence of comorbidities.

## Introduction

Despite reaching relatively high Covid-19 vaccine coverage, during the second half of 2021 many countries faced a new wave of SARS-CoV-2 infections. It may be related to : i) a variable proportion of the population not yet immunised, ii) anti-Spike antibody levels decrease over time after two doses of either RNA or ChAdOx vaccines^1^, and iii) vaccination being less efficient against the delta (B.1.617.2) variant of concern (VOC), which effectively became the single circulating form of SARS-CoV-2 during early summer 2021, compared to the original strain ^2^. Several studies have characterised the rate of such breakthrough infections in vaccinated persons ^3^, and reported that this risk increased over time after primary vaccination ^4–6^. It has also been shown that even with the delta variant, in the general population, the rate of Covid-19-related hospitalisations ^7,8^, intensive care unit (ICU) admissions, and mortality ^9^ were much lower in those who received two doses of the aforementioned vaccines.

Nevertheless, no precise data are currently available regarding the characteristics of vaccinated patients hospitalised with a SARS-CoV-2 infection. We therefore aimed to determine whether vaccination, among other factors, influenced the outcome of hospitalised patients with a SARS-CoV-2 infection, in particular in terms of need for oxygen, ICU admission, and death within 28 days after hospitalisation.

## Methods

### Population

We conducted a retrospective, multicentre study across 10 hospitals in France (Ajaccio, Annecy, Caen, Clamart, Grenoble, Le Mans, Limoges, Nancy, Nantes and Poitiers). Hospitalised patients with a PCR-proven SARS-CoV-2 infection from July to November 2021, i.e., when the Delta variant was virtually the only circulating SARS-CoV-2 variant in France, either at the time of admission or after admission, were selected. Among them, two populations were identified and included in the study:

- all patients who had received at least either one injection of the Janssen vaccine or two injections of the AstraZeneca and/or RNA vaccines (with the second dose given at least 14 days before date of PCR);
- the equivalent number of patients without any vaccination before hospital admission. This last group was composed by including for each study month and in each hospital as many unvaccinated patients as vaccinated patients, by chronological order of hospitalisation.

All the centres screened their patients to include only the delta variant. For one centre, the inclusion was prolonged up to the December 13th, but screening guaranteed that only the delta variant was involved. The following data were collected: year of birth, gender, and pre-existing comorbidities (respiratory disease, cardiac failure, arterial hypertension, solid tumour, haematological neoplasia, and kidney failure, which was determined by the Cockcroft formula based on data collected at hospital admission); Covid-19 vaccination history; clinical data at admission and minimum Ct PCR level; maximum C-reactive protein plasmatic level; extension of lung disease on computed tomography (CT-scan (if performed); need for oxygen with duration and maximal flow; need for high-flow oxygen therapy; need for ICU admission; need for invasive mechanical ventilation; prescription of steroid, tocilizumab, and/or monoclonal antibodies; and mortality at day 28.

According to the French law on ethics, patients were informed during their hospital stay that their anonymised medical data could be used for research purposes; they were given the possibility to refuse this usage. As requested by French ethics and regulatory laws, the ethical committee of the French-speaking Society of Infectious Diseases (SPILF) (IRB00011642) gave its approval for the study (N°2022-0101), and the study was declared to the French National Commission for Informatics and Liberties (CNIL MR004: n°2224742).

### Statistical analysis

Firstly, a descriptive analysis was performed in the cohort population according to vaccination status. Qualitative variables were described as counts (percentage) and frequency distributions were compared with the Chi square test or Fisher’s exact test when appropriate. Continuous variables were expressed as median (1^rst^ quartile; 3^rd^ quartile) and differences were tested with the independent t-test for normally distributed variables or otherwise the Mann-Whitney U test. Secondly, univariate and multivariate logistic regressions based on general linear models were performed with a stepwise variable selection according to the Akaike Information Criterion^10^. Factors associated with severe forms of infection defined by three outcomes, namely requirement for oxygen, ICU admission and death at day 28, were assessed, focusing on the impact of vaccination. To assess the impact of comorbidities and vaccination, we selected all the potential risk factors for severe outcome presented in Table 2 for the multivariate model (Table 3 and supplementary Table 1). In addition, to manage Missing data Not At Random (MNAR), multivariate analysis was initially performed only on the dataset with complete cases, after which the model was applied to the full dataset ^11^. Results are presented based on the full dataset, as they were similar to the analysis restricted to the complete case dataset. As an example, missing CT-scan results could equally be due to the fact that patients were too severe to benefit from a CT scan before high flow nasal oxygen or mechanical ventilation, or to the fact that the patient was mildly ill and went back home without any oxygen requirement; in some instances, the result was missing from the patient files. Finally, a multivariate analysis was similarly carried out on the dataset of vaccinated patients only, so as to independently assess the weight of comorbidities among vaccinated patients, taking into account the influence of the time elapsed since the most recent injection. Interactions were systematically searched and mentioned. All statistical analyses were performed with R version 4.1.2 ^12^.

## Results

Nine hundred seventy-four patients were initially included; 19 were excluded for i) being under 18 years (n=4), ii) not being fully vaccinated (n=12) or iii) being infected with a VOC other than delta or hospitalised when delta variant was not the only circulating variant (n=3). The characteristics of the 955 patients finally included, among whom 474 were vaccinated and 481 were not, are detailed in Table 1.

**Table 1:** Characteristics of patients with SARS-CoV-2 infection according to vaccinal status

|  |  | Vaccinated (N=474) | Unvaccinated (N=481) | p-value |
| --- | --- | --- | --- | --- |
| Age (years) n (%) | <35y | 27 (5.70) | 97 (20.17) | <0.001 |
|  | 35-65y | 111 (23.42) | 225 (46.78) |  |
|  | >65y | 336 (70.89) | 159 (33.06) |  |
| Gender: males n (%) |  | 261 (55.06) | 223 (46.36) | 0.009 |
| Body Mass Index (kg/m<sup>2</sup>)* (*out of 658*) |  | 25.76 (22.32-31.14) | 27.20 (24.15-31.22) | 0.051 |
| Overweight (BMI>25) n (% *out of 659*) |  | 192 (56.64) | 222 (69.38) | <0.001 |
| Pre-existing comorbidities n (%) | Cardiac failure (*out of 729*) | 50 (13.66) | 29 (7.99) | 0.020 |
|  | Respiratory disease (*out of 866*) | 91 (20.73) | 54 (12.65) | 0.002 |
|  | Oxygen at home (*out of 699*) | 14 (3.97) | 5 (1.45) | 0.070 |
|  | Kidney failure (GFR<30) (*out of 652*) | 51 (15.22) | 10 (3.15) | <0.001 |
|  | Diabetes (*out of 870*) | 127 (28.73) | 76 (17.76) | <0.001 |
|  | Hypertension (*out of 871*) | 244 (55.08) | 123 (28.74) | <0.001 |
|  | Solid cancer < 3 months (*out of 712*) | 17 (4.80) | 8 (2.23) | 0.070 |
|  | Hematologic malignancy (*out of 702*) | 25 (7.06) | 6 (1.72) | <0.001 |
|  | Immunodepression (*out of 723*) | 74 (20.33) | 21 (5.85) | <0.001 |
| At least one comorbidity (*out of 760*) |  | 369 (90.44) | 306 (86.93) | 0.135 |
| Tobacco n (% *out of 692*) |  | 36 (10.32) | 49 (14.29) | 0.140 |
| Previous COVID infection n (% *out of 704*) |  | 20 (5.62) | 13 (3.70) | 0.285 |
| Time from a previous infection (days)* |  | 226 (176.5-299.0]<br>(n=14) | 69 (43.3-244.3] (n=12) | 0.076 |
| Time from last vaccine dose (days)* (*out of 467*) |  | 125 (82.0-166.5] | .. |  |
| Admission for Covid-19 (*out of 955*) |  | 281 (59.28) | 361 (75.05) | <0.001 |
| Ct (lowest value if many) (*out of 452*) |  | 26.0 (18.0-34.0] | 26.0 (21.0-33.0] | 0.741 |
| CRP (highest value) (mg/L)* (*out of 687*) |  | 87.50 (31.25-152.75] | 86.0 (34.0-145.0] | 0.682 |
| Proportion of involved lung tissue in CT scanner n (% *out of 411*) | 0-10% | 67 (36.81) | 31 (13.54) | <0.001 |
|  | 11-25% | 50 (27.47) | 57 (24.89) |  |
|  | 26-50% | 39 (21.43) | 75 (32.75) |  |
|  | 51-75% | 18 (9.89) | 49 (21.40) |  |
|  | >75% | 8 (4.40) | 17 (7.42) |  |
| Requirement for oxygen n (% *out of 768*) |  | 229 (57.54) | 270 (72.97) | <0.001 |
| Maximal oxygen therapy (L/min)* (*out of 650*) |  | 3.00 (0.00-8.75] | 6.00 (2.00-50.00] | <0.001 |
| Duration of oxygen therapy (days)* (*out of 616*) |  | 3.00 (0.00-8.00] | 6.00 (2.00-12.00] | <0.001 |
| **High-flow oxygen n (% out of 718)** |  | 49 (13·57) | 105 (29·41) | <0·001 |
| **Steroid therapy** | **prescribed n (% out of 734)** | 174 (47·15) | 250 (68·49) | <0·001 |
|  | **dose median (IQR) (out of 405)** | 6·00 (6·00-7·50] | 6·00 (6·00-9·00] | 0·110 |
|  | **duration (days) (out of 407)** | 9·00 (5·00-10·00] | 10·00 (7·00-10·00] | 0·007 |
| **Tocilizumab n (% out of 656)** |  | 21 (6·31) | 59 (18·27) | <0·001 |
| **Monoclonal Ab n (% out of 734)** |  | 27 (7·26) | 7 (1·93) | <0·001 |
| **ICU admission n (% out of 739)** |  | 60 (16·22) | 133 (36·04) | <0·001 |
| **Invasive mechanical ventilation n (% out of 719)** |  | 25 (6·91) | 59 (16·53) | <0·001 |
| **Death at day 28 n (% out of 876)** |  | 74 (16·70) | 53 (12·24) | 0·075 |
| **Death at day 28 and/or ICU admission n, (% out of 750)** |  | 116 (30·69) | 166 (44·62) | <0·001 |

Quantitative variables, identified with \* are presented as median (1<sup>st</sup> quartile - 3<sup>rd</sup> quartile]. Median (1<sup>st</sup> quartile - 3<sup>rd</sup> quartile] age in years for vaccinated and unvaccinated patients was 75·0 (63·25-84·0] vs. 55·0 (38·0-73·0] (<0·001).

**Table 2:** Risk factors associated with negative outcomes (Univariate logistic regression)

| Factors |  | Death at day 28 |  | ICU |  | Oxygen required |  |
| --- | --- | --- | --- | --- | --- | --- | --- |
|  |  | OR (IC95%) | p value | OR (IC95%) | p value | OR (IC95%) | p value |
| Age * | <35y | Ref. |  | Ref. |  | Ref. |  |
|  | 35-65y | 2.93 (0.81-18.80) | 0.157 | 5.17 (2.74-10.65) | <0.001 | 5.97 (3.66-9.9) | <0.001 |
|  | >65y | 17.91 (5.56-109.62) | <0.001 | 2.32 (1.23-4.77) | 0.014 | 6.28 (3.92-10.29) | <0.001 |
| Gender: Male |  | 1.61 (1.10-2.38) | 0.015 | 1.56 (1.12-2.19) | 0.009 | 1.48 (1.10-2.00) | 0.010 |
| Diabetes mellitus |  | 1.05 (0.67-1.61) | 0.833 | 1.04 (0.71-1.52) | 0.822 | 1.37 (0.97-1.97) | 0.079 |
| Hypertension |  | 3.32 (2.23-5.01) | <0.001 | 1.00 (0.71-1.39) | 0.978 | 1.68 (1.24-2.30) | <0.001 |
| Tobacco |  | 0.48 (0.20-1.01) | 0.077 | 0.87 (0.49-1.46) | 0.602 | 0.49 (0.31-0.78) | 0.002 |
| BMI (kg/m<sup>2</sup>) |  | 1.00 (0.96-1.03) | 0.817 | 1.06 (1.03-1.09) | <0.001 | 1.06 (1.03-1.09) | <0.001 |
| Overweight (BMI>25 kg/m<sup>2</sup>) |  | 1.17 (0.74-1.87) | 0.514 | 3.30 (2.18-5.10) | <0.001 | 2.13 (1.53-2.99) | <0.001 |
| Kidney failure GFR<30) |  | 4.55 (2.56-7.98) | <0.001 | 0.28 (0.10-0.61) | 0.004 | 1.02 (0.58-1.84) | 0.954 |
| Respiratory disease |  | 1.23 (0.75-1.96) | 0.386 | 0.94 (0.60-1.44) | 0.794 | 1.57 (1.05-2.40) | 0.031 |
| Oxygen at home |  | 2.68 (0.92-6.94) | 0.051 | 0.46 (0.07-1.67) | 0.305 | 1.96 (0.68-7.01) | 0.244 |
| Cardiac failure |  | 3.45 (2.04-5.77) | <0.001 | 0.68 (0.36-1.23) | 0.224 | 2.11 (1.22-3.80) | 0.010 |
| Solid cancer < 3 months |  | 4.03 (1.71-9.13) | <0.001 | 0.44 (0.10-1.29) | 0.183 | 0.58 (0.25-1.31) | 0.184 |
| Hematologic malignancy |  | 2.39 (1.02-5.19) | 0.034 | 0.78 (0.28-1.82) | 0.588 | 0.71 (0.34-1.49) | 0.353 |
| Immunosuppression |  | 2.23 (1.32-3.68) | 0.002 | 0.91 (0.54-1.50) | 0.723 | 1.00 (0.64-1.57) | 0.986 |
| At least one comorbidity |  | 4.43 (1.80-14.73) | 0.004 | 2.25 (1.26-4.35) | 0.010 | 2.75 (1.74-4.38) | <0.001 |
| Previous SARS-CoV-2 infection |  | 0.17 (0.02-1.28) | 0.086 | 0.10 (0.01-0.70) | 0.021 | 0.13 (0.05-0.32) | <0.001 |
| Time from last infection (days) |  | 1.03 (0.95-1.11) | 0.525 | 1.00 (0.99-1.01) | 0.835 | 1.00 (0.99-1.00) | 0.436 |
| Vaccination |  | 1.44 (0.98-2.11) | 0.062 | 0.34 (0.24-0.48) | <0.001 | 0.50 (0.37-0.68) | <0.001 |
| Time from last vaccine injection (days) |  | 1.01 (1.00-1.01) | <0.001 | 1.00 (0.99-1.00) | 0.341 | 1.01 (1.00-1.01) | <0.001 |

\*Results were similar for age as a continuous variable, as it was associated to death at day 28 with an OR (IC95%) at 1.06 (1.05-1.08) and a p-value <0.001, to ICU with an OR (IC95%) at 0.99 (0.98-1.00) and a p-value at 0.049 and to Oxygen requirement with an OR (IC95%) at 1.02 (1.02-1.03) and a p-value at <0.001.

**Table 3:**
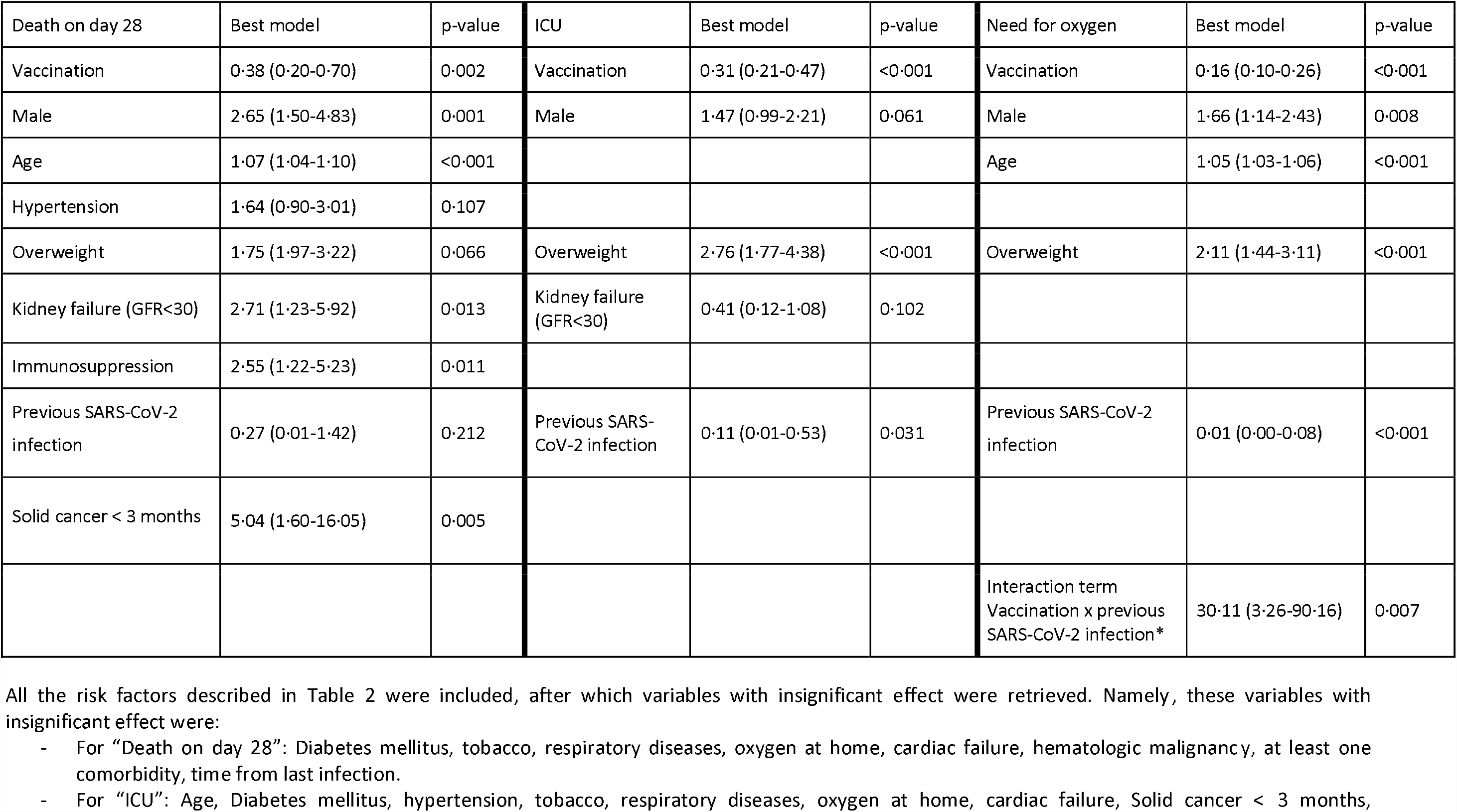
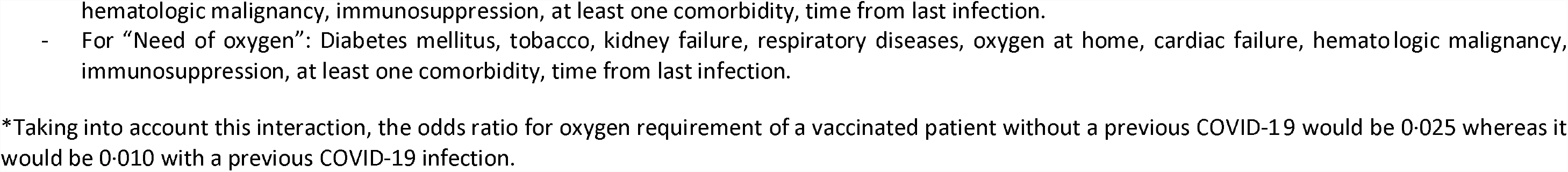
Factors associated with negative outcomes, with a multivariate logistic regression (best model according to AIC).

| Death on day 28 | Best model | p-value | ICU | Best model | p-value | Need for oxygen | Best model | p-value |
| --- | --- | --- | --- | --- | --- | --- | --- | --- |
| Vaccination | 0.38 (0.20-0.70) | 0.002 | Vaccination | 0.31 (0.21-0.47) | <0.001 | Vaccination | 0.16 (0.10-0.26) | <0.001 |
| Male | 2.65 (1.50-4.83) | 0.001 | Male | 1.47 (0.99-2.21) | 0.061 | Male | 1.66 (1.14-2.43) | 0.008 |
| Age | 1.07 (1.04-1.10) | <0.001 |  |  |  | Age | 1.05 (1.03-1.06) | <0.001 |
| Hypertension | 1.64 (0.90-3.01) | 0.107 |  |  |  |  |  |  |
| Overweight | 1.75 (1.97-3.22) | 0.066 | Overweight | 2.76 (1.77-4.38) | <0.001 | Overweight | 2.11 (1.44-3.11) | <0.001 |
| Kidney failure (GFR<30) | 2.71 (1.23-5.92) | 0.013 | Kidney failure (GFR<30) | 0.41 (0.12-1.08) | 0.102 |  |  |  |
| Immunosuppression | 2.55 (1.22-5.23) | 0.011 |  |  |  |  |  |  |
| Previous SARS-CoV-2 infection | 0.27 (0.01-1.42) | 0.212 | Previous SARS-CoV-2 infection | 0.11 (0.01-0.53) | 0.031 | Previous SARS-CoV-2 infection | 0.01 (0.00-0.08) | <0.001 |
| Solid cancer < 3 months | 5.04 (1.60-16.05) | 0.005 |  |  |  |  |  |  |
|  |  |  |  |  |  | Interaction term<br>Vaccination x previous SARS-CoV-2 infection* | 30.11 (3.26-90.16) | 0.007 |

- For “Death on day 28”: Diabetes mellitus, tobacco, respiratory diseases, oxygen at home, cardiac failure, hematologic malignancy, at least one comorbidity, time from last infection. - For “ICU”: Age, Diabetes mellitus, hypertension, tobacco, respiratory diseases, oxygen at home, cardiac failure, Solid cancer < 3 months,
hematologic malignancy, immunosuppression, at least one comorbidity, time from last infection.
- For "Need of oxygen": Diabetes mellitus, tobacco, kidney failure, respiratory diseases, oxygen at home, cardiac failure, hematologic malignancy, immunosuppression, at least one comorbidity, time from last infection.
\*Taking into account this interaction, the odds ratio for oxygen requirement of a vaccinated patient without a previous COVID-19 would be 0.025 whereas it would be 0.010 with a previous COVID-19 infection.

The two groups differed on several points: unvaccinated patients were significantly younger and had fewer comorbidities (reflecting the sequential prioritisation of the vaccine campaign during the early months of 2021), although they also had a higher body mass index. Among vaccinated patients, the median delay since the most recent dose was 125 days. The BNT162b2 (Comirnaty, Pfizer) vaccine had been used in 357 (76·1%) patients. Astra-Zeneca, Janssen (Johnson & Johnson), Moderna and Sputnik had been used in 62 (13·2%), 27 (5·8%), 22 (4·7%) and 1 (0·2%) patients respectively.

All in all, vaccinated patients had milder forms of SARS-CoV-2 infection (Table 1). They were less frequently hospitalised because of the infection itself; they had fewer extended lung lesions on CT-scan; they required oxygen therapy less frequently, with a lower oxygen flow, and for a shorter duration; they received steroid therapy less often; they were less frequently admitted to ICU; and they less often needed high-flow oxygen or mechanical ventilation. Vaccinated patients’ 28-day survival was not statistically different, even in patients presenting more comorbidities; in addition, when considering a negative outcome index combining death at day 28 and ICU requirement, there was a significant relative 32% risk reduction among vaccinated patients compared to unvaccinated patients. The protective effect of vaccination was also observed when considering only older patients (>65 years), as illustrated in Figure 1.

**Figure 1:**
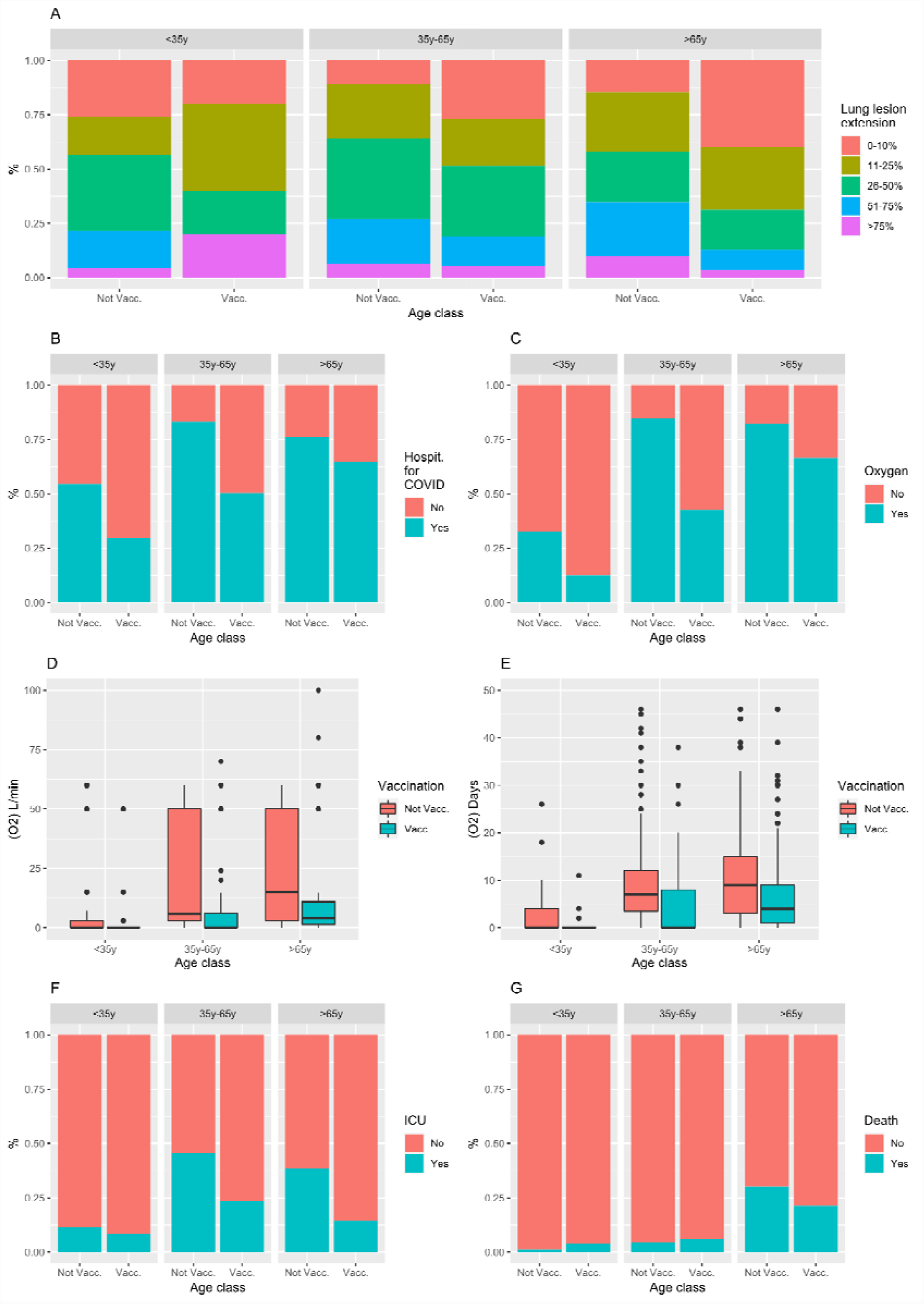
SARS-CoV-2 infection severity criteria according to vaccine status in the three age classes. A: Extension of lung lesions on CT scanner; B: hospitalisation cause; C: Proportion of patients requiring oxygen; D: Maximal oxygen flow; E: Duration of oxygen therapy; F: Proportion of patients requiring ICU and G: Proportion of death at day 28.

In univariate analysis, vaccination and a previous SARS-CoV-2 infection were protective toward ICU and oxygen requirement, but not significantly associated with 28-day mortality. Comorbidities, older age, male gender and a longer time lapse since the most recent vaccine injection (among vaccinated patients) were significantly associated with a negative outcome (Table 2). In addition, being hospitalised for COVID, an elevated CRP, a low Cycle threshold and extensive lung lesions on CT scanner were associated with a negative outcome (Supplementary Table 2).

Multivariate analysis confirmed the protective effect of vaccination regarding the need for oxygen, the need for ICU admission, and the risk of death. Vaccination was even able to offset the impact of many risk factors such as hypertension and overweight (Table 3 and Supplementary Table 1). Notably, in addition to factors with odds ratio not statistically different from 1 presented in Table 3, many risk factors were finally not kept in the final multivariate model due to their negligible impact. Moreover, in the multivariate regression model for death at day 28, the absence of significant interactions also showed that the protective effect of vaccination occurs for all patients, including those most at risk for severe forms such as older or immunosuppressed patients. In the multivariate regression model for oxygen requirement, an interaction between vaccination and a previous infection by COVID-19 was observed, suggesting that a previous COVID-19 infection decreased the influence of the vaccination; the large coefficient associated with having previous COVID-19 infection suggests that it takes away most of the protective effect on oxygen requirement, leaving a small but still significant influence to vaccination.

## Discussion

The emergence of SARS-CoV-2 delta VOC jeopardised resolution of the Covid-19 pandemic. Indeed, the humoral immune response triggered by natural infection and/or vaccination is less likely to neutralise the VOC, which acquired key mutations in gene coding for the Spike protein. We therefore aimed to characterise the protection associated with vaccination towards the consequences of Delta VOC infection by studying the population of patients hospitalised with this infection. Including both patients hospitalised because of Covid-19 and those with incidental positive PCR while being admitted for another purpose allowed us to compare vaccinated and non-vaccinated patients not only with Covid-19, but all infected patients, including those who were asymptomatic. Indeed, vaccine-induced protection can manifest by the absence of symptoms in case of infection ^13^.

The emergence of the Delta variant was anticipated as being associated with breakthrough infections. Indeed, *in vitro experiments* showed that the neutralisation index of plasma from vaccinated individuals was lower for delta than for initial viral strain ^14–16^. Different studies and a meta-analysis assessing the impact of primary vaccination observed a ≃ 10% reduction in vaccine efficacy against asymptomatic and symptomatic infections with delta VOC, but no reduction of vaccine efficacy against severe forms ^2,17^. Some authors have noted that the higher incidence of infections with this VOC may also be due to the fact that when delta became predominant, those infected had received their primary vaccination 4 to 10 months before, meaning that the infection may have been related to the waning of the level of vaccine-induced antibodies ^18^.

Recent studies showed 86-88% vaccine-associated reduction in the hospital admission rate in those infected with the delta VOC ^19–21^; Our observations confirm the positive impact of vaccination on the severity of the SARS-CoV-2 delta infection, even in hospitalised patients. Moreover, we observed that the positive impact was maintained in populations at risk of severe Covid-19: those aged over 65 years, and those with pre-existing cardiac failure, kidney failure, or a chronic respiratory disease (at least for one or several severity criteria). All in all, this legitimates the prioritisation of these populations during the early months of the 2021 vaccine campaign: they benefited from valuable protection, even with the delta variant. Taken with previous fatality studies of vaccinated patients ^21–23^, this also confirms the perception shared by many physicians that vaccinated fragile people survived a form of Covid-19 in 2021 that they would not have survived without vaccination. Taking into account that overweight was more frequent among unvaccinated patients, while vaccination nonetheless effectively protected the associated mortality, emphasis should be made on this risk factor for severe forms of Covid-19.

The association between vaccination and less severe SARS-CoV-2 infection was less clearly evidenced in immunocompromised patients. The absence of interaction between vaccination and immunosuppression and the absence of a significant effect of immunosuppression in the model applied specifically to vaccinated patients suggest that vaccination is also efficient in immunocompromised patients (suppl. Table 1). Nonetheless, these results should not be over-interpreted as most of the immunocompromised patients were vaccinated in our cohort, and there is probably a lack of power to detect an interaction between both variables, or in other words a diminished effect of vaccination in immunocompromised patients. Moreover, immunocompromised patients remained at high risk of a severe form (OR=2·55) even after adjustment for parameters such as other risk factors and vaccination (Table 1). Decreased efficacy in this population was previously noted for virtually all vaccines, particularly influenza vaccines ^24^, and was also observed with Covid-19 vaccines ^25^. This led some countries to recommend an additional dose of the vaccine to these patients, and the prophylactic use of passive immunisation by monoclonal antibodies in immunocompromised patients with no or insufficient antibody response to vaccines ^26^.

As older patients were not only at risk of severe forms, but were also vaccinated earlier in the vaccination campaign, a lack of protection may have existed due to the aforementioned waning of neutralising antibody titres over time after vaccination. The multivariate analysis we specifically applied to vaccinated patients showed that both age and time from vaccination were associated with a significant increase in 28-day mortality. Once again, this confirms the need for a booster vaccine injection, particularly in those most at risk.

Our study has several limitations. Firstly, due to the retrospective design of the study, some data retrieved from the medical files may have been erroneous or not completely accurate. However, the high number of studied patients, and comprehensive analysis on patients of known risk factors for adverse outcomes helped to reduce risk of biases. Secondly, we did not measure anti-Spike antibody titers in vaccinated patients at admission, a previously described predictive risk factor for SARS-CoV-2 infection and Covid-19 ^27^; it would have helped to better understand whether severe infections in vaccinated subjects were due to waning of the neutralising antibody vaccine response. Moreover, we used ICU admission as a severity criterion; however, it is usually considered that older patients with many comorbidities would not benefit from ICU admission in terms of survival. Therefore, not being admitted to ICU does not necessarily mean that there is no severe form of Covid-19.

In conclusion, patients hospitalised with a SARS-CoV-2 infection with the delta VOC have less marked severity criteria when they are vaccinated more than 14 days before hospitalisation, especially in case of elderly patients. These data should be provided when communicating information about Covid-19 vaccination with subjects at risk of severe Covid-19 because of their age or comorbidities.

## Data Availability

All data produced in the present study are available upon reasonable request to the authors

## Supplementary materials

**Supplementary table 1:** Factors associated with negative outcomes, with a multivariate logistic regression applied to vaccinated patients only (best model according to AIC).

| Death on day 28 | Best model |  | ICU admission | Best model |  | Need for oxygen | Best model |  |
| --- | --- | --- | --- | --- | --- | --- | --- | --- |
|  | OR (IC95%) | p value |  | OR (IC95%) | p value |  | OR (IC95%) | p value |
| Male | 3.39 [1.53; 8.17] | **0.004** |  |  |  |  |  |  |
| Age | 1.04 [1.01; 1.08] | **0.034** |  |  |  | Age | 1.03 [1.02; 1.05] | **<0.001** |
| Overweight | 2.28 [1.07; 5.10] | 0.037 | Overweight | 2.68 [1.35; 5.65] | **0.006** | Overweight | 1.79 [1.08; 3.00] | **0.026** |
| Kidney failure (GFR<30) | 3.87 [1.60; 9.46] | 0.003 | Kidney failure (GFR<30) | 0.36 [0.08; 1.08] | 0.107 |  |  |  |
|  |  |  |  |  |  | Respiratory disease | 1.53 [0.85; 2.81] | 0.165 |
|  |  |  |  |  |  | Tobacco | 0.32 [0.12; 0.76] | **0.013** |
| Solid cancer < 3 months | 13.28 [3.51; 52.36] | **<0.001** |  |  |  |  |  |  |
| Last Vaccine delay | 1.01 [1.00; 1.02] | 0.014 |  |  |  |  |  |  |
|  |  |  |  |  |  | Preexisting COVID | 0.39 [0.12; 1.16] | 0.104 |
|  |  |  |  |  |  | Cardiac failure | 1.81 [0.85; 4.39] | 0.167 |

**Supplementary table 2:** Characteristics of COVID-19 associated with negative outcomes (Univariate logistic regression)

| Factors | Death at day 28 |  | ICU |  | Oxygen required |  |
| --- | --- | --- | --- | --- | --- | --- |
|  | OR (IC95%) | p value | OR (IC95%) | p value | OR (IC95%) | p value |
| Hospitalised for COVID-19 | 1.54 [1.03; 2.37] | **0.041** | 6.52 [4.26; 10.31] | **<0.001** | 18.42 [12.71; 27.12] | **<0.001** |
| CRP (highest value if many) | 1.01 [1.00; 1.01] | **<0.001** | 1.00 [1.00; 1.01] | **<0.001** | 1.01 [1.01; 1.01] | **<0.001** |
| Ct value | 0.92 [0.89; 0.95] | **<0.001** | 0.96 [0.93; 0.99] | **0.005** | 0.90 [0.87; 0.92] | **<0.001** |
| Proportion of involved lung tissue in CT scanner |  |  |  |  |  |  |
| 0-10% | .. |  | .. |  | .. |  |
| 11-25% | 1.08 [0.47; 2.50] | 0.859 | 2.08 [0.94; 4.85] | 0.078 | .. |  |
| 26-50% | 0.99 [0.43; 2.29] | 0.984 | 9.18 [4.49; 20.48] | **<0.001** | .. |  |
| 51-75% | 2.22 [0.98; 5.17] | 0.058 | 24.71 [10.91; 61.04] | **<0.001** | .. |  |
| >75% | 4.72 [1.72; 13.02] | **0.002** | 61.60 [17.71; 296.29] | **<0.001** | .. |  |

## References

1 Goldberg Y, Mandel M, Bar-On YM, et al. Waning Immunity after the BNT162b2 Vaccine in Israel. N Engl J Med 2021; 385: e85.

2 Harder T, Külper-Schiek W, Reda S, et al. Effectiveness of COVID-19 vaccines against SARS-CoV-2 infection with the Delta (B.1.617.2) variant: second interim results of a living systematic review and meta-analysis, 1 January to 25 August 2021. Euro Surveill Bull Eur Sur Mal Transm Eur Commun Dis Bull 2021; 26. DOI:10.2807/1560-7917.ES.2021.26.41.2100920.

3 Keehner J, Horton LE, Binkin NJ, et al. Resurgence of SARS-CoV-2 Infection in a Highly Vaccinated Health System Workforce. N Engl J Med 2021; 385: 1330–2.

4 Khoury J, Najjar-Debbiny R, Hanna A, et al. COVID-19 vaccine - Long term immune decline and breakthrough infections. Vaccine 2021; 39: 6984–9.

5 Mizrahi B, Lotan R, Kalkstein N, et al. Correlation of SARS-CoV-2-breakthrough infections to time-from-vaccine. Nat Commun 2021; 12: 6379.

6 Lebourgeois S, Menidjel R, Chenane HR, et al. Alpha (B.1.1.7) and Delta (B.1.617.2 - AY.40) SARS-CoV-2 variants present strong neutralization decay at M4 post-vaccination and a faster replication rates than D614G (B.1) lineage. J Infect 2021; : S0163-4453(21)00556-9.

7 Grannis SJ, Rowley EA, Ong TC, et al. Interim Estimates of COVID-19 Vaccine Effectiveness Against COVID-19-Associated Emergency Department or Urgent Care Clinic Encounters and Hospitalizations Among Adults During SARS-CoV-2 B.1.617.2 (Delta) Variant Predominance - Nine States, June-August 2021. MMWR Morb Mortal Wkly Rep 2021; 70: 1291–3.

8 Harris JE. COVID-19 Incidence and Hospitalization During the Delta Surge Were Inversely Related to Vaccination Coverage Among the Most Populous U.S. Counties. Health Policy Technol 2021; : 100583.

9 Direction-de-la-recherche-des-études-de-l’évaluation-et-des-statistiques. (2021, novembre). Neuf fois plus d’entrées en soins critiques parmi les personnes non vaccinées que parmi celles qui sont complètement vaccinées de 20 ans et plus. Ministère Santé Solidar. 2021. https://drees.solidarites-sante.gouv.fr/communique-de-presse/neuf-fois-plus-dentrees-en-soins-critiques-parmi-les-personnes-non-vaccinees. (accessed Dec 23, 2021).

10 Akaike H. Information theory and an extension of the maximum likelihood principle. In: 2nd International Symposium on Information Theory. Academiai Kiado, 1973.

11 Jakobsen JC, Gluud C, Wetterslev J, Winkel P. When and how should multiple imputation be used for handling missing data in randomised clinical trials – a practical guide with flowcharts. BMC Med Res Methodol 2017; 17: 162.

12 R Development Core Team. R: A language and environment for statistical computing. R Foundation for Statistical Computing, Vienna, Austria. ISBN 3-900051-07-0. 2012. http://www.R-project.org/.

13 Rovida F, Cassaniti I, Paolucci S, et al. SARS-CoV-2 vaccine breakthrough infections with the alpha variant are asymptomatic or mildly symptomatic among health care workers. Nat Commun 2021; 12: 6032.

14 Luczkowiak J, Labiod N, Rivas G, et al. Neutralizing response against SARS-CoV-2 variants 8 months after BNT162b2 vaccination in naïve and COVID-19 convalescent individuals. J Infect Dis 2021; : jiab634.

15 Lustig Y, Zuckerman N, Nemet I, et al. Neutralising capacity against Delta (B.1.617.2) and other variants of concern following Comirnaty (BNT162b2, BioNTech/Pfizer) vaccination in health care workers, Israel. Euro Surveill Bull Eur Sur Mal Transm Eur Commun Dis Bull 2021; 26. DOI:10.2807/1560-7917.ES.2021.26.26.2100557.

16 Davis C, Logan N, Tyson G, et al. Reduced neutralisation of the Delta (B.1.617.2) SARS-CoV-2 variant of concern following vaccination. PLoS Pathog 2021; 17: e1010022.

17 Grant R, Charmet T, Schaeffer L, et al. Impact of SARS-CoV-2 Delta variant on incubation, transmission settings and vaccine effectiveness: Results from a nationwide case-control study in France. Lancet Reg Health Eur 2021; : 100278.

18 Bosch W, Cowart JB, Bhakta S, et al. COVID-19 Vaccine-Breakthrough Infections Requiring Hospitalization in Mayo Clinic Florida through August 2021. Clin Infect Dis Off Publ Infect Dis Soc Am 2021; : ciab932.

19 Poukka E, Baum U, Palmu AA, et al. Cohort study of Covid-19 vaccine effectiveness among healthcare workers in Finland, December 2020 - October 2021. Vaccine 2021; : S0264-410X(21)01640-6.

20 Bruxvoort KJ, Sy LS, Qian L, et al. Effectiveness of mRNA-1273 against delta, mu, and other emerging variants of SARS-CoV-2: test negative case-control study. BMJ 2021; 375: e068848.

21 Katikireddi SV, Cerqueira-Silva T, Vasileiou E, et al. Two-dose ChAdOx1 nCoV-19 vaccine protection against COVID-19 hospital admissions and deaths over time: a retrospective, population-based cohort study in Scotland and Brazil. Lancet Lond Engl 2022; 399: 25–35.

22 Tang P, Hasan MR, Chemaitelly H, et al. BNT162b2 and mRNA-1273 COVID-19 vaccine effectiveness against the SARS-CoV-2 Delta variant in Qatar. Nat Med 2021; 27: 2136–43.

23 Arbel R, Hammerman A, Sergienko R, et al. BNT162b2 Vaccine Booster and Mortality Due to Covid-19. N Engl J Med 2021; 385: 2413–20.

24 Beck CR, McKenzie BC, Hashim AB, Harris RC, University of Nottingham Influenza and the ImmunoCompromised (UNIIC) Study Group,, Nguyen-Van-Tam JS. Influenza vaccination for immunocompromised patients: systematic review and meta-analysis by etiology. J Infect Dis 2012; 206: 1250–9.

25 Marra AR, Kobayashi T, Suzuki H, et al. Short-term effectiveness of COVID-19 vaccines in immunocompromised patients: A systematic literature review and meta-analysis. J Infect 2022; : S0163-4453(21)00658-7.

26 EMEA. Ronapreve (casirivimab and imdevimab) An overview of Ronapreve and why it is authorised in the EU. 2021. https://www.ema.europa.eu/documents/overview/ronapreve-epar-medicine-overview_en.pdf (accessed Jan 16, 2022).

27 Khoury DS, Cromer D, Reynaldi A, et al. Neutralizing antibody levels are highly predictive of immune protection from symptomatic SARS-CoV-2 infection. Nat Med 2021; 27: 1205–11.

